# Details of COVID-19 Disease Mitigation Strategies in 17 K-12 Schools in Wood County, Wisconsin

**DOI:** 10.1101/2021.03.16.21253761

**Authors:** Amy Falk, Alison Benda, Peter Falk, Sarah Steffen, Mikaela DeCoster, Monica Gandhi, Tracy Beth Høeg

**Affiliations:** Department of Pediatrics, Aspirus Doctors Clinic, 2031 Peach St. Wisconsin Rapids, WI; Medical College of Wisconsin - Central Wisconsin, Wausau, WI; ReVision Eye Care, Wisconsin Rapids WI; Center for AIDS Research, Division of HIV, Infectious Diseases, and Global Medicine, Department of Medicine, University of California, San Francisco, CA; Department of Physical Medicine & Rehabilitation, University of California - Davis; Northern California Orthopaedic Associates, Grass Valley, CA

**Author notes:** Corresponding author: Amy Falk, MD, Work/personal.

## Abstract

**Importance:** With the current COVID-19 return-to-school guidelines, over half of America’s K-12 students are being denied access to full time in-person education, leading to harmful academic, emotional and health consequences.

**Objective:** To describe the specific details of mitigation strategies employed at 17 K-12 schools in Wisconsin during a time of exceptionally high COVID-19 community disease prevalence where in-school transmission was minimal. The aim of this report is to assist school districts and governing bodies in developing full-time return to school plans.

**Design:** Retrospective cohort

**Setting:** Wood County, Wisconsin, August 31–November 29, 2020

**Participants:** 5,530 students and staff from 17 schools in 4 school districts

**Main outcomes and measures:**

1. Distancing between primary and secondary students in school
2. School ventilation details
3. Masking among teachers
4. Lunch, recess and bussing practices

**Results:** 89.3% of elementary students included in our study did not maintain 6 feet of physical distancing in the classroom and 94.8% were within 6 feet in lunchrooms. The majority of secondary students (86.2%) were able to maintain 6 feet of distancing in the classroom but no students were greater than 6 feet in the hallways. 58.8% of schools did not install new ventilation systems prior to the school year. Students ate lunch indoors. Bussing of students continued and all elementary children were allowed to go without masks at recess.

**Conclusion and relevance:** In the setting of high community COVID-19 disease transmission, 6 feet of distance between elementary students and major ventilation system renovations in primary or secondary schools do not appear to be necessary to minimize disease spread. Requiring masks at recess and prohibiting bussing also appears unnecessary. These findings may inform guidance on the safe reopening of schools and allow for more children to return to in-person schooling.

## Introduction

Since March of 2020, approximately 1/3 of children in our country’s schools have had no in-person learning due to the COVID-19 pandemic *(1)* and over a half continue to not be in school full time (2). An estimated 3 million K-12 students haven’t engaged in any form of on-line learning and are considered lost to the educational system since the start of the pandemic *(3)*. There are growing concerns about the harms of prolonged school closures. On January 26th, 2021, our group’s study (4) “COVID-19 Cases and Transmission in 17 K–12 Schools — Wood County, Wisconsin, August 31–November 29, 2020” published in the CDC’s journal, *MMWR*, reported low in-school disease transmission rates in a community with a test positivity rate of up to 41.6% and up to 1,189/100,000 weekly cases. Of the 191 cases identified in schools during the 13 week study period, 7 cases (5 elementary, 2 secondary students) were determined to be contracted in school. Zero cases were contracted by staff in school. These findings have been cited *(5)* as evidence that K-12 schools can safely reopen despite high community disease spread. On February 26th, 2021, the CDC released Operational Guidelines (6) advising against full-time in-person learning for elementary students or any in-person learning for secondary students when community transmission is above 10% test positivity or 100 cases/100,000 per week. For reference, our study had community disease rates up to 12 times higher than this threshold and our school openings were successful.

Furthermore, the new guidelines recommend students maintain at least 6 feet of distance with community transmission 24 times lower than in our study (50/100,000 weekly cases). The space required to achieve this amount of distancing is resulting in many schools remaining closed or in hybrid mode. Recent data on student and staff rates of COVID-19 among classes with students seated 3 or 6 feet apart found no significant differences in overall disease incidence. (7). The purpose of this report is to provide further detail on distancing among students in class and at lunch in a setting of high community disease spread and minimal in-school transmission. We also report on school ventilation changes, recess policies, bussing practices and masking among teachers. Further information is also provided on the circumstances of in-school transmission of cases.

## Methods

From August 31, 2020 to November 29, 2020, COVID-19 transmission data and masking compliance rates were collected from 8 elementary and 9 secondary schools with a total population of 5,530 students and teachers. In February 2021, a follow-up Google Forms survey was distributed to administration at each of the 17 participating schools. The questionnaire gathered information about distancing between students, teacher masking compliance, recess practices, and ventilation. The protocol was reviewed by the Aspirus Wausau Hospital Institutional Review Board and determined to be exempt from human subjects review as it met the requirements under 45 CFR 46. 104 (d) (2) and underwent a limited review as required under 46.111(a)(7).

### Elementary Schools

This cohort included 1,529 elementary students and 275 staff. In classrooms, 90.7% of students were seated less than 6 feet from each other and 9.3% distancing 6 feet or more (Figure 1; Table 1). During lunch, only 5.2% of students were spaced 6 feet or more with the remaining 94.8% sitting less than 6 feet apart. In hallways 85.2% of students were 3-6 feet apart and the remaining were less than 3 feet. At recess 85.6% of children did not wear masks. All elementary students were allowed to utilize outdoor playground equipment and other recess toys.

**Figure 1.**
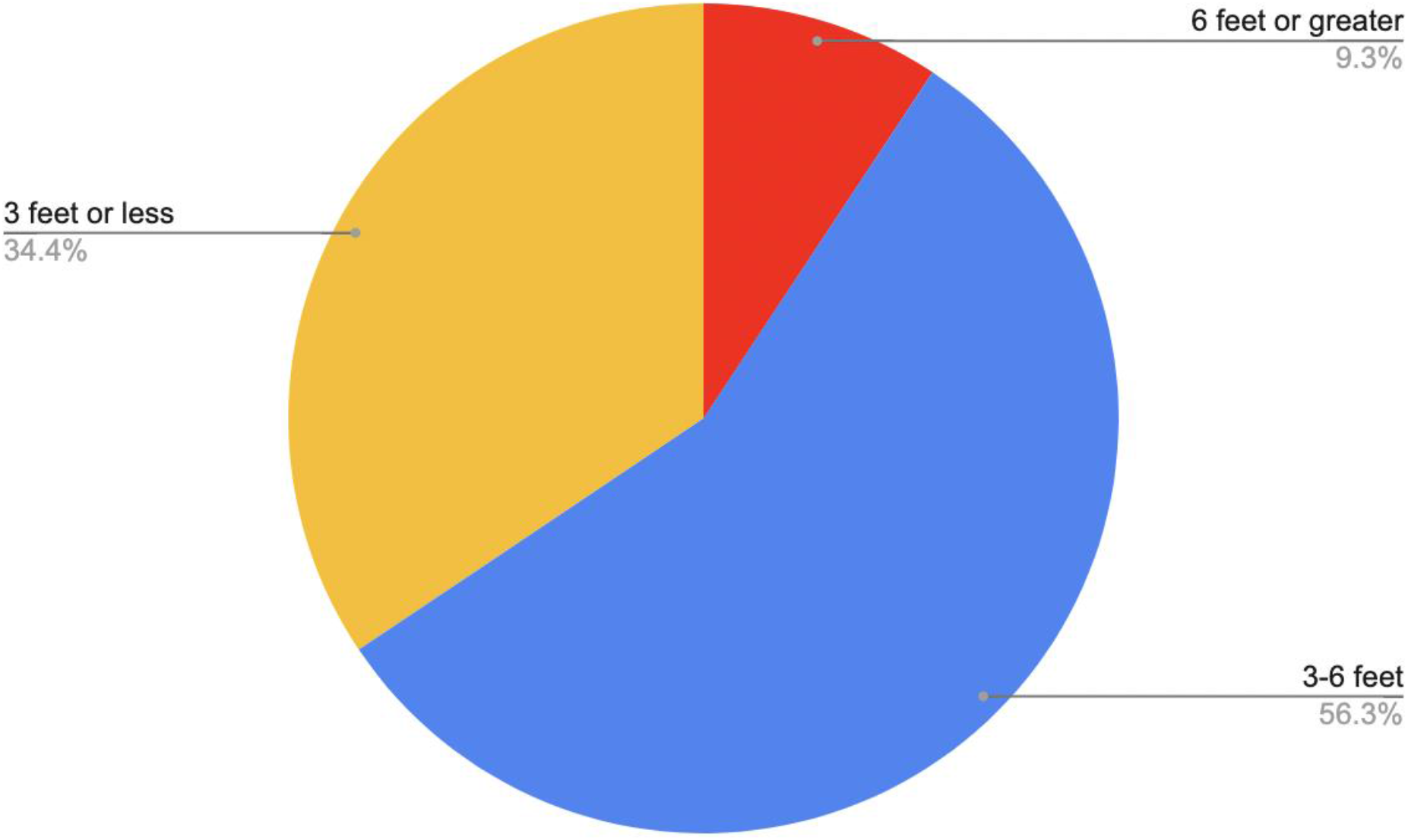
Distance between students in primary classrooms.

**Table 1.** Mitigation Strategies Among Students and in Schools

|  | *Elementary n*<br>*(%)* | *Secondary n*<br>*(%)* |
| --- | --- | --- |
| ***Distancing in the classroom*** |  |  |
| < 3 feet between students | 526 (34.7) | 227 (6.8) |
| 3 to 6 feet between students | 861 (56.0) | 235 (7) |
| > 6 feet between students | 142 (9.3) | 2885 (86.2) |
| ***Distancing at lunch*** |  |  |
| 6' or more | 1450 (94.8) | 2280 (68.1) |
| <6 feet | 79 (5.2) | 1067 (31.9) |
| ***The students eat lunch...*** |  |  |
| Cafeteria only | 1252 (81.9) | 2806 (83.8) |
| Cafeteria and other rooms | 277 (18.1) | 541 (16.2) |
| ***Distancing in hallways*** |  |  |
| 0-3' between students | 227 (14.8) | 1901 (56.8) |
| 3-6' between students | 1302 (85.2) | 1446 (43.2) |
| ***Children wear masks at recess*** |  |  |
| Yes | 142 (9.3) | NA |
| No | 1308 (85.6) | NA |
| ***New School Ventilation System*** |  |  |
| Yes | 34.6 (3) | 57.2 (4) |
| No | 65.4 (5) | 42.8 (5) |
| ***Windows Regularly Opened*** |  |  |
| Yes | 25.3 (2) | 0 |
| No | 74.7 (6) | 100 (9) |

### Secondary Schools

This cohort included 3,347 secondary students and 379 staff. In classrooms, 86.2% of secondary students maintained at least 6 feet of distance, 7.0% maintained between 3 and 6 feet and 6.8% less than 3 feet (Figure 2; Table 1). At lunch, 68.1% of students were seated greater than 6 feet apart and the remaining 31.9% were less than 6 feet apart. In the hallways, 56.8% of students were frequently between 0-3 feet, while the remainder were typically between 3-6 feet.

**Figure 2.**
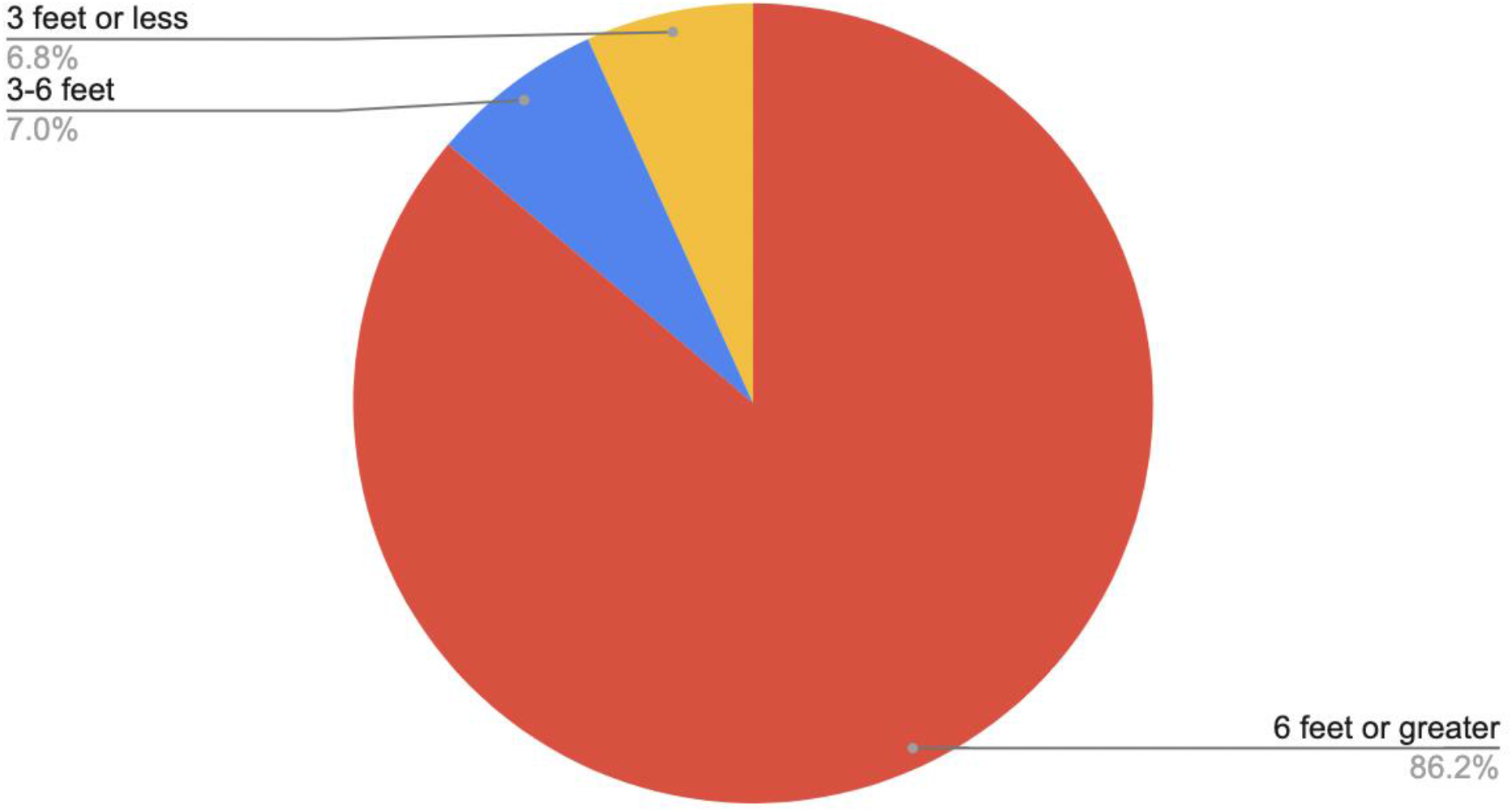
Distance between students in secondary classrooms.

### Teachers

Masking compliance among teachers while indoors was 90% or greater at all elementary and 7/9 secondary schools. 2/9 secondary schools, which had 6.9% of the secondary student population, reported teachers were masked 75-90% of the time.

### Ventilation and bussing

41.2% of schools installed new air filtration systems that affected the entire school before the Fall 2020 school year; the remaining 58.8% did not (Figure 3, Table1). Regular opening of windows occurred in 11.8% of schools (Figure 4, Table 1).

**Figure 3.**
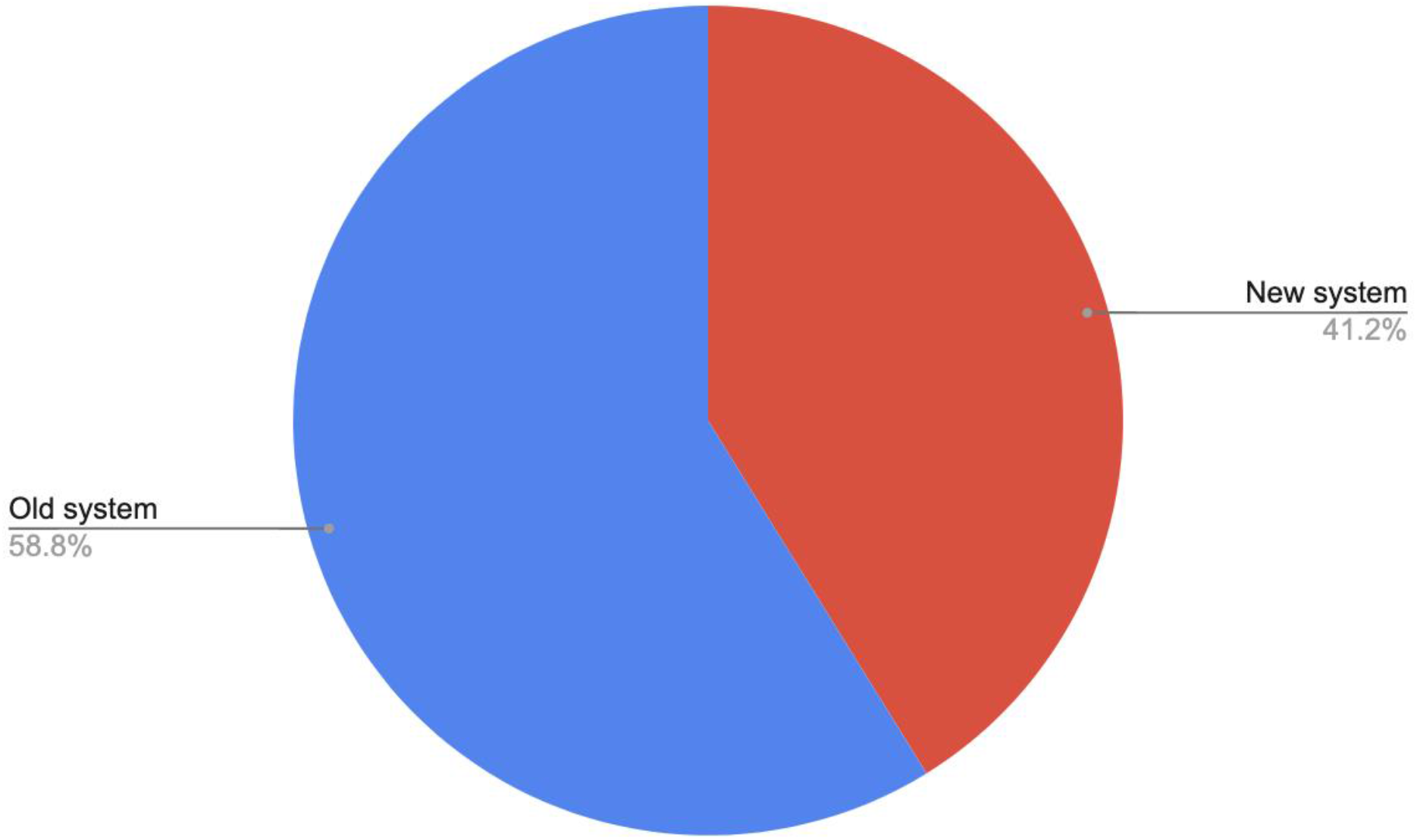
School ventilation systems.

**Figure 4.**
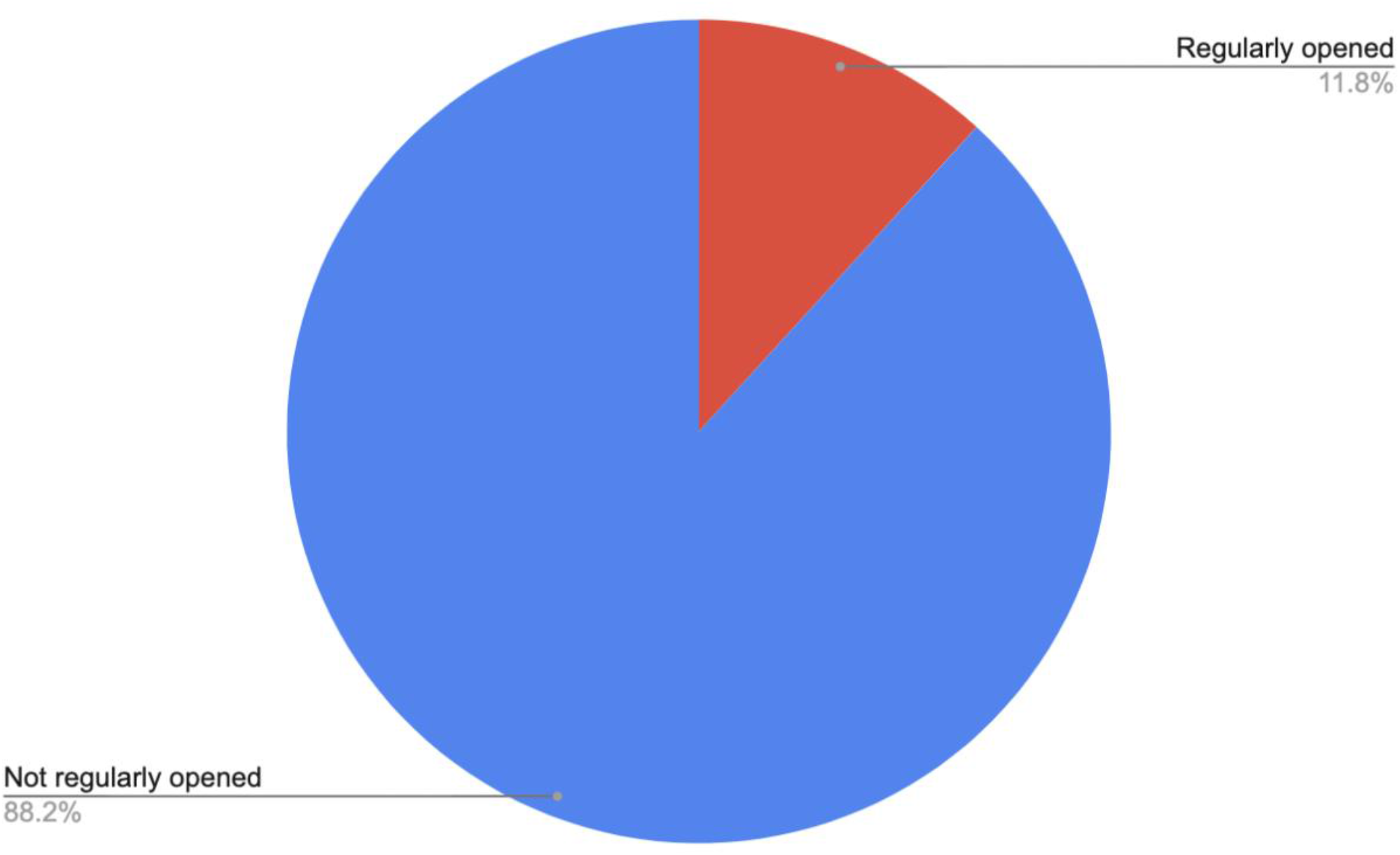
Classroom window opening practices.

All schools offered bussing for students. Masking was required of students and drivers. Physical distancing was attempted on busses, but seating children together was reported. Schools reported 25-50% of students utilized bussing services.

### In-school disease spread and distancing of students

There were 7 total COVID-19 cases in children (none in staff) attributed to in-school spread: five elementary students and two secondary students. Three elementary cases occurred in one school which maintained 3-6 feet of distance in the classroom and greater than 6 feet at lunchtime. The remaining elementary cases occurred at separate schools; one school maintaining greater than 6 feet between students in the classroom and the other maintaining 0-3 feet. Both schools utilized greater than 6 feet distance at lunch. The two cases of COVID-19 among secondary students occurred at separate schools. One school described 3-6 feet between students; the other had greater than 6 feet in the classroom. At lunch, both schools maintained greater than 6 feet distancing. Due to the minimal amount of in-school spread in our study, it is not possible to rule out a correlation between distancing and disease spread.

## Discussion

COVID-19 disease mitigation strategies in this K-12 population across 17 schools in Wisconsin varied greatly. Minimal in-school disease transmission was reported despite a community test positivity rate between 7 and 41.6% in the 13-week study period. During this time, the vast majority of elementary schools did not maintain 6 feet between students in classroom cohorts, while the majority, but not all, of secondary schools did. These results suggest that maintaining 6 feet distance between students, particularly not at the elementary level, is not required to effectively mitigate in-school SARS-CoV-2 spread.

At recess, most students did not mask or distance and were allowed to play with playground equipment within their cohorts. This has important implications for the well-being of children. Recess can provide necessary physical activity as well as a break from masking, which may make it easier for children to resume required indoor mitigation strategies.

Opening windows to increase airflow was not commonly done. Approximately half the schools changed air filtration systems without any obvious detriment to those that didn’t. This might reassure school districts where these mitigation measures are not structurally or financially possible.

Within the range of mitigation strategies outlined above, including nearly-universal indoor masking, cohorting and distancing as much as possible, there was minimal transmission among students and none to staff. Despite high community disease prevalence at the time of our study, stricter measures than those outlined in this report do not appear necessary for schools to safely reopen.

## Data Availability

All data reported in this publication are available upon request through the corresponding author.

## Acknowledgements

The school districts of south Wood County, Craig Broeren, Sue Kunferman, Dr. Lisa Olson. The authors report no relevant conflicts of interest.

